# Directed functional brain connectivity is altered in sub-threshold amyloid-β accumulators

**DOI:** 10.1101/2022.10.26.22281539

**Authors:** Mite Mijalkov, Dániel Veréb, Anna Canal-Garcia, Giovanni Volpe, Joana B. Pereira, Alzheimer’s Disease Neuroimaging Initiative

## Abstract

Several studies have shown that amyloid-β (Aβ) deposition below the clinically relevant cut-off levels is associated with subtle changes in cognitive function and increases the risk for developing future Alzheimer’s disease (AD). Although functional MRI is sensitive to early alterations occurring during AD, sub-threshold changes in Aβ levels have not been linked to functional connectivity measures. This study aimed to apply directed functional connectivity to identify early changes in network function in cognitively unimpaired participants who, at baseline, exhibit Aβ accumulation below the clinically relevant threshold. To this end, we analyzed baseline functional MRI data from 113 cognitively unimpaired participants of the Alzheimer’s Disease Neuroimaging Initiative cohort who underwent at least one ^18^F-florbetapir-PET after the baseline scan. Using the longitudinal PET data, we classified these participants as Aβ negative (Aβ−) non-accumulators (n=46) and Aβ-accumulators (n=31). We also included 36 individuals who were amyloid-positive (Aβ+) at baseline and continued to accumulate Aβ (Aβ+ accumulators). For each participant, we calculated whole-brain directed functional connectivity networks using our own anti-symmetric correlation method and evaluated their global and nodal properties using measures of network segregation (clustering coefficient) and integration (global efficiency). When compared to Aβ-non-accumulators, the Aβ-accumulators showed lower global clustering coefficient. Moreover, the Aβ+ accumulator group exhibited reduced global efficiency and clustering coefficient, which at the nodal level mainly affected the superior frontal gyrus, anterior cingulate cortex and caudate nucleus. In Aβ-accumulators, global measures were associated with lower baseline regional PET uptake values, as well as higher scores on the Modified Preclinical Alzheimer Cognitive Composite. Our findings indicate that directed connectivity network properties are sensitive to subtle changes occurring in individuals who have not yet reached the threshold for Aβ positivity, which makes them a potentially viable marker to detect negative downstream effects of very early Aβ pathology.

**Significance Statement:**

- Directed functional connectivity measures are sensitive to very early changes in amyloid-β deposition.
- Amyloid-β burden is associated with decreases in global and nodal directed measures of network integration and segregation.
- Measures of directed network integration and segregation are associated with subclinical cognitive changes in Aβ-negative accumulators.

## Introduction

Alzheimer’s disease (AD) is a neurodegenerative disorder that results in a progressive loss of memory and other cognitive functions^1^. However, the pathological processes in AD begin many years before any symptoms of dementia and other cognitive impairments become apparent^2^. In particular, Aβ accumulation, which can be measured by an increased burden in amyloid positron emission tomography (PET), is an early sign of preclinical AD stages, being one of the first events in the sequence of pathological processes that ultimately results in the development of dementia^3,4^.

Although Aβ plaques are a hallmark of AD, studies using amyloid PET imaging found they are also present in the brains of otherwise healthy older individuals who show no indications of cognitive impairment^5, 6^. The deposition of Aβ in such individuals occurs over a very long time period^7^ and results in changes in the functional connectivity patterns^8^. However, widespread Aβ changes may not be necessary for observing connectivity changes ^9^. Instead, such effects can be evident also in the presence of emergent Aβ pathology^9^, the process of amyloid accumulation in cognitively healthy people who have not yet reached the threshold for clinical Aβ positivity. Such emergent pathology has been associated with higher tau accumulation and atrophy rates, as well as memory impairment and alterations in functional connectivity in previous studies^10–14^.

Here, we assessed whether early Aβ accumulation in otherwise healthy, cognitively normal individuals that are Aβ-negative at baseline has an effect on the organization of whole-brain directed functional connectivity networks. The directed functional networks were calculated using the method of anti-symmetric delayed correlations that we developed earlier^15^ since this method is more sensitive to detect subtle changes in functional connectivity when compared to conventional methods. Our results show that early amyloid deposition is associated with a decrease of segregation in functional networks. These changes were also observed, although to a greater extent, in individuals that were already Aβ+ at baseline, who additionally showed changes in functional integration. Moreover, these network properties were associated with the extent of amyloid burden and preclinical cognitive composite scores. These findings suggest that, even at sub-threshold Aβ deposition levels, the decrease of directional flow in the functional network could be an indication of neuronal changes and subtle changes in cognition in individuals at the preclinical stages of AD.

## Materials & Methods

### Participants

Data used in the preparation of this article were obtained from the Alzheimer’s Disease Neuroimaging Initiative (ADNI) database (adni.loni.usc.edu). The ADNI was launched in 2003 as a public-private partnership, led by Principal Investigator Michael W. Weiner, MD. The primary goal of ADNI has been to test whether serial magnetic resonance imaging (MRI), PET, other biological markers, and clinical and neuropsychological assessment can be combined to measure the progression of mild cognitive impairment (MCI) and early AD. ADNI is conducted in accordance with the ethical standards of the institutional research committees and with the 1975 Helsinki declaration and its later amendments. Written informed consent, obtained from all subjects and/or authorized representatives and study partners, and ethical permits have been obtained at each participating site of ADNI and we have signed the data user agreements to analyze the data.

For the purposes of this study, only cognitively normal participants from ADNI3 were included with baseline functional MRI and at least two ^18^F-Florbetapir PET scans. The inclusion/exclusion criteria for ADNI are described in detail at http://www.adni-info.org/. In short, participants were aged between 55 and 90 years, were fluent in Spanish or English while having also completed at least 6 years of education, had Clinical Dementia Rating scores of 0 and no other significant neurological disorder. We used the Preclinical Alzheimer Cognitive Composite^16^ (PACC, with trails test) to assess the cognitive status of the individuals. This composite calculates a composite score based on tests that assess global cognition, episodic memory and executive function and has been shown to be sensitive to the very initial signs of cognitive decline, before the appearance of clinical symptoms of mild cognitive impairment stage.

### Image Acquisition and Preprocessing

Subjects in the ADNI3 study underwent standardized MRI scanning protocols at the acquisition sites using 3T MRI scanners from different vendors, and 18F-florbetapir PET scans. The full list of MRI/PET scanners and detailed imaging protocols can be found at the ADNI study website (https://adni.loni.usc.edu/). From the full suite of imaging data, we used the sagittal 3D accelerated MPRAGE sequence (full head coverage, voxel size = 1×1×1 mm^3^, field of view = 208×240×256 mm^3^, repetition time = 2300 ms, inversion time = 900 ms) and the axial EPI-BOLD functional MRI sequence (voxel size = 3.4×3.4×3.4 mm^3^, field of view = 220×220×163 mm^3^, repetition time = 3000 ms, echo time = 30 ms, flip angle = 90°). Functional and structural MRI scans were pre-processed using a standardized pipeline implemented in fMRIPrep^17^(v20.2.4, https://fmriprep.org/en/stable/). The resulting functional images additionally underwent motion correction using the Friston-24 head motion model^18^ and nuisance regression for signal from the white matter and cerebrospinal fluid. 18F-florbetapir PET scans were acquired in 4×5 minute frames, 50-70 minutes after the injection of 10 mCi dose on average.

### Group classification

We divided the participants into 3 groups based on Aβ levels measured by ^18^F-Florbetapir PET. Using their baseline scan as a reference, we classified the participants as being Aβ-negative or Aβ-positive using the standardized uptake value ratios (SUVRs) for a global composite region that included the caudal anterior cingulate, frontal, lateral parietal and lateral temporal gyri on 18F-Florbetapir PET, normalized by the whole cerebellum. A previously defined cut-off was used to determine amyloid-positivity^19^. Using the longitudinal composite-normalized cortical summary SUVR values, we further subdivided the Aβ-negative individuals into two groups that showed either positive (accumulators) annual rate of change in their Aβ levels (i.e., the change in the SUVR values per year) or remained largely stable with a small negative rate of change in Aβ levels (non-accumulators). While the negative slopes could represent measurement noise, some studies have suggested that there is a possibility that they are due to Aβ clearance^14^. This procedure resulted in 3 groups of participants: 31 Aβ-negative non-accumulators (Aβ−/ NAc); 46 Aβ-negative accumulators (Aβ−/ Ac) and 36 Aβ-positive (Aβ+). The characteristics of each group are summarized in Table 1, and they were compared between all groups using the Kruskal–Wallis rank sum test.

**Table 1:** Demographic and clinical data of the participants.

|  | **Aβ- Non-accumulators**<br>**(n = 46)** | **Aβ- Accumulators**<br>**(n = 31)** | **Aβ+ Accumulators**<br>**(n = 36)** | **P values** |
| --- | --- | --- | --- | --- |
| **Age** | 75.57 (6.24) | 76.26 (6.35) | 78.27 (6.32) | 0.112 |
| **Sex (M / F)** | 23 / 23 | 10 / 21 | 19 / 17 | 0.19 |
| **Education** | 16.78 (2.30) | 16.71 (2.51) | 16.17 (2.59) | 0.482 |
| **APOE ε4 carriers (%)** | 15.22% | 30% | 45.71% | 0.007 |
| **Aβ PET composite SUVR (baseline)** | 0.66 (0.11) | 0.69 (0.19) | 0.87 (0.12) | < 0.001 |
| **Rate of change in Aβ PET composite SUVR** | -0.0121 (0.0096) | 0.0179 (0.0159) | 0.0344 (0.0190) | < 0.001 |
| **MMSE** | 28.93 (1.39) | 29.42 (0.89) | 28.67 (1.64) | 0.053 |
| **PACCtrailsB** | 0.42 (1.89) | 0.17 (2.41) | -0.73 (2.89) | 0.219 |

Means are followed by standard deviations. Aβ , amyloid-β ; Aβ -, Aβ-negative; Aβ +, Aβ-positive; M / F, Male / Female; APOE $\epsilon$ 4, apolipoprotein E $\epsilon$ 4 allele; PET SUVR, positron emission tomography standardized uptake value ratios; MMSE, Mini Mental State Examination; PACCtrailsB, modified Preclinical Alzheimer Cognitive Composite with trails test.

### Anti-symmetric functional networks capture directed functional connectivity between brain areas

Temporal delays can arise in the interaction between brain areas because of the dispersed nature of brain regions and the limited speed of information transfer between them^20^. Therefore, to achieve a coherent characterization of the functional connectivity, it is crucial to capture the information contained in this complicated temporal lag structure^21,22^. To this purpose, we have developed an anti-symmetric correlation-based approach for determining the directed connections between pairs of brain areas, allowing for the computation of whole-brain directed functional connectivity networks^15^. In this approach, if one brain area’s activation time series has similar properties to the time-shifted version of the activation pattern in another brain region, then the first region is considered to have a directed interaction with the second region. The level of directed interaction is quantified by the inverse of the time lag between the time series of those regions (which captures the fact that directly connected brain regions are expected to activate with a much shorter delay that indirectly connected regions). The direction of the interaction is determined by the order of precedence in time (i.e., the early region is the source, and the late region is the end of the connection).

Figure 1A illustrates the calculation of the anti-symmetric connectivity networks for a set of five brain regions and their corresponding time series. The lagged correlation matrix can be evaluated by calculating the time-lagged Pearson’s correlation coefficient between all pairs of regions at a given temporal lag. This matrix captures the directed connection between the regions in both directions, i.e. the matrix entries *(i, j)* and *(j, i)* provide an estimate from the directed connection from region *i* to *j* and vice versa. As any other square matrix, the lagged correlation adjacency matrix can be uniquely expressed as the sum of a symmetric and anti-symmetric matrix. We use the anti-symmetric matrix to approximate the whole-brain directed functional connectivity; this method summarizes the directed connection between regions *I* and *j* with a single entry which captures its direction and the magnitude. More details about the method are presented elsewhere^15^.

**Figure 1:**
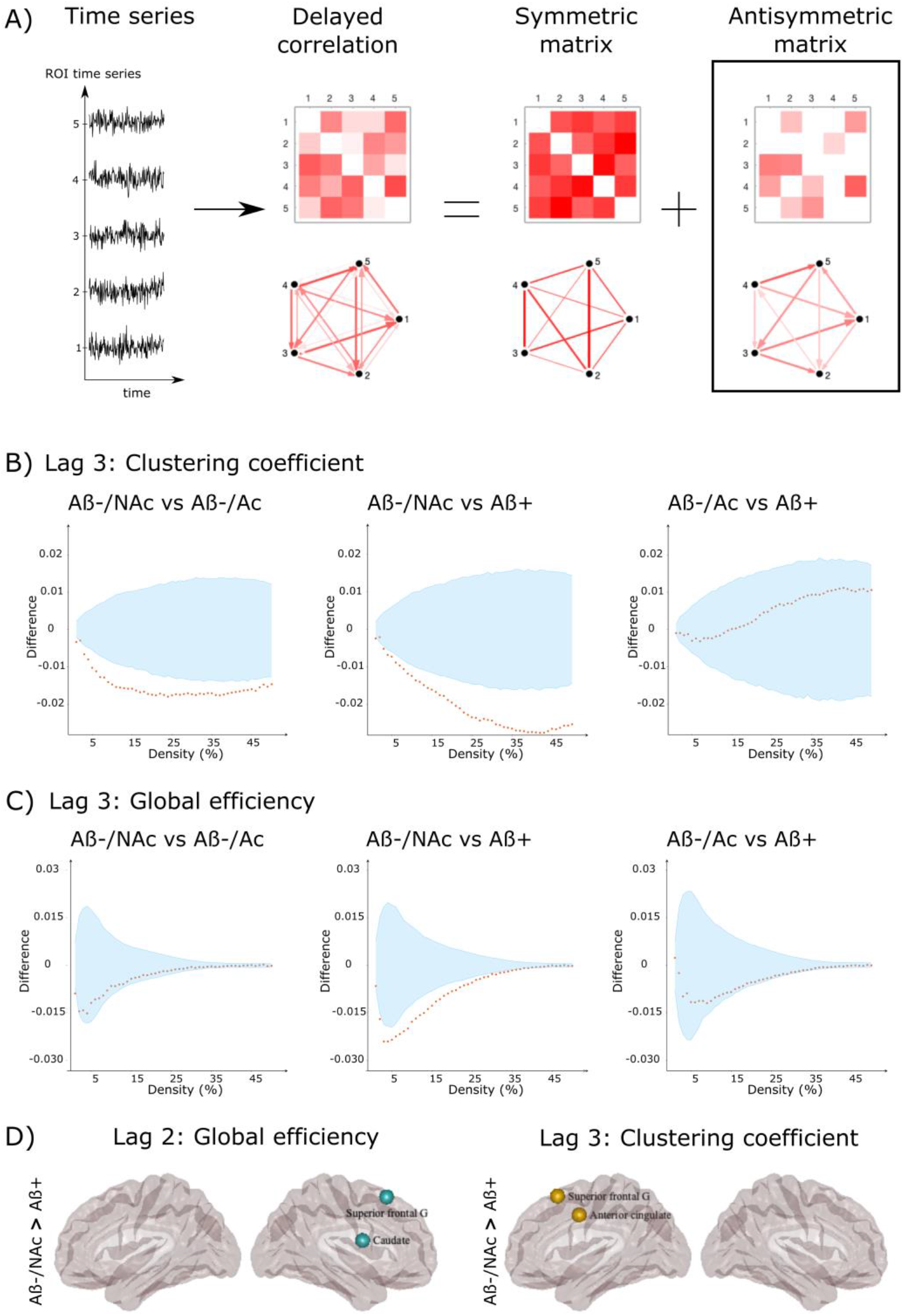
Sample Differences between different groups in global and nodal directed functional network topology. A) Overview of the calculation procedure for the anti-symmetric correlation. For simplicity, we illustrate the procedure using the time activation series for five nodes as an example. The lagged Pearson’s correlation coefficient between these time series at varying temporal lags allows the estimation of lagged correlation functional networks (in this example, at lag of 1). The lagged adjacency matrix can be written as a sum of symmetric and anti-symmetric and matrices. Here, we use the anti-symmetric matrices to estimate the functional connectivity of each individual. Differences between the different groups at lag 3 (calculated as group 2 - group 1 in the figure) in the B) clustering coefficient and C) global efficiency. The differences in the corresponding network measures between groups are plotted in orange circles as a function of network density and the upper and lower bounds of the 95% confidence intervals (CI) are plotted in blue. The differences are considered statistically significant if they fall outside the CIs. D) Differences between Aβ−/ NAc and Aβ+ groups in nodal network measures. The plotted regions remained significant after correcting for multiple comparisons using FDR at q < 0.05.

### Network construction and analysis

We calculated a 200 × 200 weighted connectivity matrix for each participant, with the edges generated using anti-symmetric correlation and the nodes corresponding to the 200 Craddock atlas brain regions^23^. We calculated a binary matrix for each adjacency matrix in which the correlation coefficient was assigned a value of 1 if it was greater than a certain threshold and 0 otherwise. We performed this binarization procedure throughout the complete range of network densities (*D*) available to the anti-symmetric correlation (D_range_ = 1% −50%) and compared the global topology of the binary networks across that range. In the case of nodal measures, we estimated measure-specific area under the curve (AUC) by numerically integrating the nodal measure values over the whole density range; the curves depict the evolution of the appropriate nodal network measure as a function of the network density for a specific brain area. These AUC values were used to assess the differences in the nodal directed connectivity patterns between the different groups. We assessed the global and nodal topology by calculating directed clustering coefficient and global efficiency using the BRAPH software package^24^.

### Statistical analysis

The statistical significance of differences in network measures across groups was determined using nonparametric permutation testing with 10000 permutations, with p < 0.05 considered significant for a two-tailed test of the null hypothesis. The nodal measures were corrected for multiple comparisons by using the Benjamini-Hochberg procedure^25^ to apply false discovery rate (FDR) corrections at q < 0.05 to adjust for the number of brain regions. All analysis were carried out using age, sex, and education as covariates. The correlation analyses were performed at each density for the global measures and for brain regions that showed between-group significant differences. All network measures were adjusted for age, sex, and education before carrying out the correlation analyses. In the case of global measures, the correlations were considered consistent if it remained significant at extended range of densities after the application of FDR correction to control for the number of densities.

## Results

### Amyloid deposition results in changes of functional segregation and integration properties of the network

We have previously shown that the changes in directed functional connectivity due to neurodegenerative processes can be captured by small temporal lags, up to the lag of 7^15^. As the current study employs functional MRI scans that have been acquired with longer repetition times, we evaluate the between-group differences at small delays up to lag of 5, which corresponds to the same temporal scale used in ref. 15. The between-group differences in the clustering coefficient and global efficiency at lag of 3 and are shown in Figure 1B and 1C respectively, whereas the corresponding results for lags 2 and 4 are plotted in Supplementary Fig. S1. The anti-symmetric correlation methods showed widespread significant decreases in the clustering coefficient in the Aβ-negative accumulators when compared to Aβ-negative non-accumulators group. Similar differences were observed in the Aβ+ group; in addition, the Aβ+ group showed also decreased global efficiency when compared to Aβ-negative non-accumulators (Fig. 1B and 1C). The differences between Aβ+ individuals and Aβ-non-accumulators were mostly pronounced in superior frontal gyrus, caudate nucleus and anterior cingulate cortex at different temporal lags (Fig. 1D).

### Directed functional connectivity is associated with global amyloid deposition level and cognitive scores in Aβ-accumulators

In the Aβ-accumulators group, the clustering coefficient was significantly associated with baseline global SUVR values at lag 4 at high density values (33% −50%). The global efficiency showed significant associations with the SUVR values at lags 2 (5%-9% and 13% to 39%), 3 (3% to 40%) and 4 (2% to 39%). Regarding the cognitive tests, the PACC with trails test scores showed significant association with the global efficiency at lag 2 (18% to 34%). A representative correlation for each association at a single density is shown in Figure 2.

**Figure 2:**
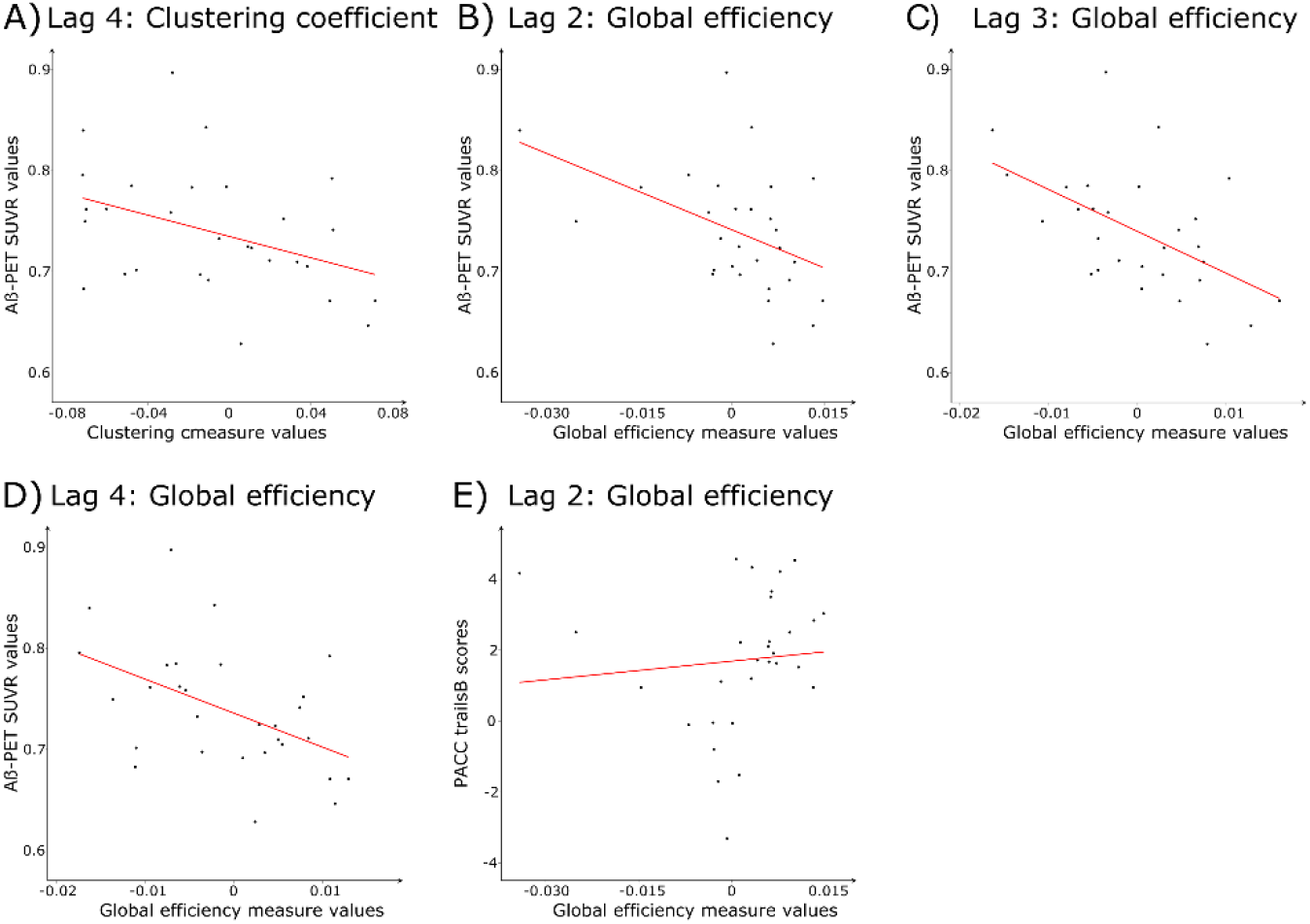
Correlation between Aβ deposition levels and cognitive scores and global measures for Aβ−/ Ac group. The plots show the best line fit for Aβ−/ Ac group between baseline Aβ SUVR deposition levels and A) clustering coefficient at lag 4, and global efficiency at lags B) 2, C) 3, D)4. The correlation between PACCtrailsB test scores and global efficiency at lag 2 is shown in F). The global measure values were regressed with age, sex, and education as covariates; the residuals of the regression were used to calculate the best line fit. A representative fit at density A) 39% and B-F) 25% is shown.

## Discussion

Anti-symmetric correlations can be used for quantifying directed connectivity between brain regions by harvesting the information contained in the temporal lags between the activation time series of different regions^15^. In this study, we used this approach to demonstrate the impact of Aβ deposition on directed functional connectivity in a group of healthy, cognitively normal adults. We observed that groups with larger Aβ load exhibited reduced levels of functional segregation and integration in their functional connectomes, both at global network levels as well as in the superior frontal gyrus, caudate nucleus and anterior cingulate cortex. The global measures in the Aβ negative group with positive rates of amyloid accumulation were associated with global Aβ load and cognitive tests, indicating that they are sensitive to pathological and clinical alterations that may occur during preclinical AD. Overall, our results suggest that the directional flow in brain activity includes unique information that might serve as a novel biomarker to study functional abnormalities in preclinical AD.

In the preclinical stage of AD, accumulating protein aggregates already affect neurons and induce synaptic dysfunction that can be detected with fMRI even in the absence of express neurodegeneration^26^. In particular, before the presence of detectable amyloid plaques in the brain, soluble Aβ oligomers already appear in the intracellular space and disrupt neurotransmission on both pre- and postsynaptic levels^27^, while exerting a specific effect on glutamatergic transmission by interfering with the mechanisms of long-term depression and potentiation that contribute to early memory deficits during the course of the disease^28^. The earliest known sites of Aβ-accumulation are the precuneus, posterior cingulate and medial prefrontal cortices, areas that comprise the core of the default mode network^28^. Accordingly, functional connectivity measures (especially those of the default mode network) are sensitive to these early changes and were shown to correlate with pre- and postsynaptic markers in amyloid-positive individuals^30^. Individuals that demonstrate a trajectory of accumulating amyloid-levels still below the defined clinically relevant threshold also exhibit functional network reorganization, mainly in the default mode and salience networks^31^. Our results are in line with these reports and additionally show that directed approaches are more sensitive to changes due to subthreshold amyloid accumulation than conventional methods.

Anti-symmetric correlation networks had an abnormal global topology in the individuals with higher Aβ deposition, characterized by decreases in global efficiency and clustering coefficient. When compared to Aβ-non-accumulators, these differences were more pronounced in the Aβ positive group than in Aβ negative accumulators, suggesting a link between the topology and amount of Aβ in the brain. The clustering coefficient provides an estimation of the functional segregation of the connectivity network and the ability of performing specialized tasks between neighboring regions, while the global efficiency estimates the functional integration and efficiency of the information transfer between distant brain regions^24, 32^. The existence of locally clustered connections and high functional integration are indicative of a small-world network, an organization characteristic of a well-functioning brain network^32, 33^. Therefore, the reductions in the clustering coefficient and global efficiency can be interpreted as a loss of the small-world property of the functional networks, a finding consistently observed in AD^8, 34, 35^. Given the importance of functional connectivity for cognitive performance^36^, the disruptions in the integrity of the functional networks can have a significant role in the development of cognitive deficits in AD. This is also supported by our findings, which demonstrate that impaired cognitive function is linked to dysfunctional functional integration in preclinical AD.

Regarding alterations at the nodal level, we found reduced global efficiency and clustering coefficient in the superior frontal gyrus, the anterior cingulate cortex and the caudate nucleus in Aβ positive individuals. The superior frontal gyrus is part of the frontoparietal executive and control networks important in executive function^37^. The disruption of network measures in the superior frontal gyrus is in line with previous reports showing that executive function is affected early on in the disease course^38^. The anterior cingulate cortex is a prominent area of the salience network that is also impaired in AD^31^. Functional changes of the caudate nucleus are not widely reported in AD, however, a loss of volume in the caudate nucleus has been reported as an early sign of the disease^39^. Additionally, these areas participate in the function of the default mode network, one of the earliest affected brain networks in AD^40^. To summarize, alterations of nodal network characteristics reinforce earlier reports of executive dysfunction as one of the earliest affected cognitive domains during the course of AD, while also affecting the default mode network.

In conclusion, we show that directed functional connectivity networks are altered in sub-threshold Aβ-accumulators. The properties of these networks are associated with levels of amyloid-deposition and cognitive test scores reflecting executive function, which potentiates their use as a possible marker for early amyloid-related pathology.

## Data Availability

Data used in the preparation of this article were obtained from the Alzheimer's Disease Neuroimaging Initiative (ADNI) database (adni.loni.usc.edu).

## Acknowledgements

Data collection and sharing of ADNI was funded by the National Institutes of Health Grant U01 AG024904 and Department of Defense award number W81XWH-12-2-0012. ADNI is funded by the National Institute on Aging, the National Institute of Biomedical Imaging and Bioengineering, and through generous contributions from the following: Alzheimer’s Association; Alzheimer’s Drug Discovery Foundation; BioClinica, Inc.; Biogen Idec Inc.; Bristol-Myers Squibb Company; Eisai Inc.; Elan Pharmaceuticals, Inc.; Eli Lilly and Company; F. Hoffmann-LaRoche Ltd and its affiliated company Genentech, Inc.; GEHealthcare; Innogenetics, N.V.; IXICO Ltd; Janssen Alzheimer Immunotherapy Research \& Development, LLC.; Johnson \& Johnson Pharmaceutical Research \& Development LLC.; Medpace, Inc.; Merck \& Co., Inc.; Meso Scale Diagnostics, LLC.; NeuroRx Research; Novartis Pharmaceuticals Corporation; Pfizer Inc.; Piramal Imaging; Servier; Synarc Inc.; and Takeda Pharmaceutical Company. The Canadian Institutes of Health Research is providing funds to support ADNI clinical sites in Canada. Private sector contributions are facilitated by the Foundation for the National Institutes of Health (www.fnih.org). The grantee organization is the Northern California Institute for Research and Education, and the study is coordinated by the Alzheimer’s Disease Cooperative Study at the University of California, San Diego. ADNI data are disseminated by the Laboratory for Neuro Imaging at the University of California, Los Angeles.

