## Supplementary Information for "Directed functional brain connectivity is altered in sub-threshold amyloid-β accumulators"

**Mite Mijalkov[1,*], Dániel Veréb[1], Anna Canal-Garcia[1], Giovanni Volpe[2], Joana B. Pereira[1,3,*], Alzheimer's Disease Neuroimaging Initiative**

[1] Department of Neurobiology, Care Sciences and Society, Karolinska Institutet, Stockholm, Sweden.

[2] Department of Physics, Goteborg University, Gotebörg, Sweden.

[3] Memory Research Unit, Department of Clinical Sciences Malmö, Lund University, Lund, Sweden.

* Corresponding authors: Mite Mijalkov and Joana B. Pereira,

Address: KI, Dept. NVS, division of clinical geriatrics, Neo 7th floor, Blickagången 16, 141 83 Huddinge, Sweden.

\ //

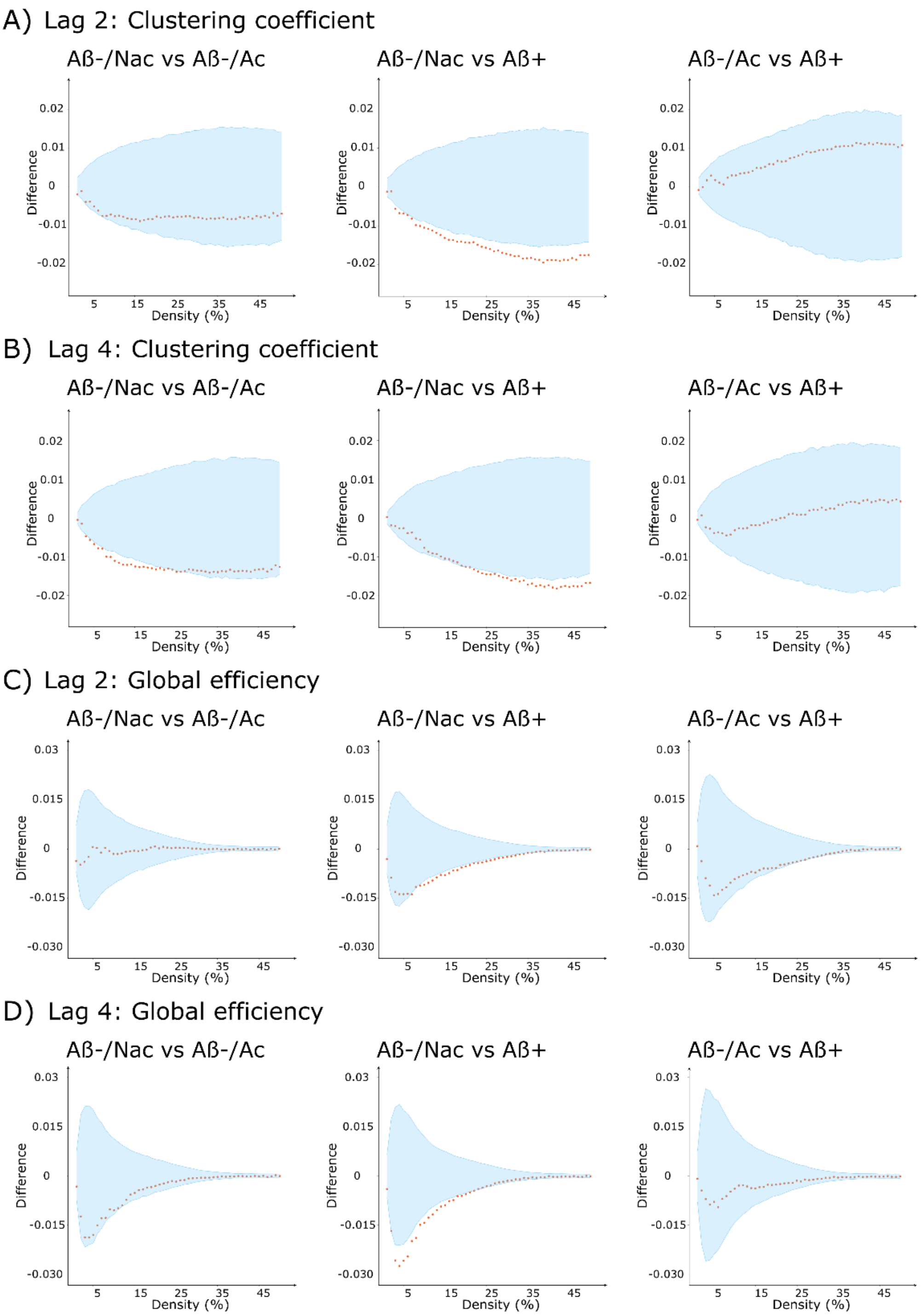


**Fig S1. Differences between different groups in global directed functional network topology.** Between-group differences (calculated as group 2 - group 1 in the figure) in clustering coefficient at A) lag 2 and B) lag 4 as well as in global efficiency at C) lag 2 and D) lag 4. Orange circles represent the differences in the corresponding network measures as a function of network density; the upper and lower bounds of the 95% confidence intervals (CI) are plotted in blue. Differences are considered statistically significant if they fall outside the CIs.
